# Development of a Novel Serological Assay for the Detection of Mpox Infection in Vaccinated Populations

**DOI:** 10.1101/2023.04.18.23288419

**Authors:** Jennifer L. Yates, Danielle T. Hunt, Karen E. Kulas, Karen Chave, Linda Styer, Sandhya T. Chakravarthi, Gianna Y. Cai, Maria C. Bermúdez-González, Giulio Kleiner, Deena Altman, Komal Srivastava, PVI study group, Viviana Simon, Dennis Feihel, Joseph McGowan, Wayne Hogrefe, Philip Noone, Christina Egan, Mark K. Slifka, William T. Lee

## Abstract

In 2022 the World Health Organization declared a Public Health Emergency for an outbreak of mpox, the zoonotic Orthopoxvirus (OPV) affecting at least 103 non-endemic locations world-wide. Serologic detection of mpox infection is problematic, however, due to considerable antigenic and serologic cross-reactivity among OPVs and smallpox-vaccinated individuals. In this report, we developed a high-throughput multiplex microsphere immunoassay (MIA) using a combination of mpox-specific peptides and cross-reactive OPV proteins that results in the specific serologic detection of mpox infection with 93% sensitivity and 98% specificity. The New York State Non-Vaccinia Orthopoxvirus Microsphere Immunoassay is an important diagnostic tool to detect subclinical mpox infection and understand the extent of mpox spread in the community through retrospective analysis.

## Introduction

Mpox (formerly known as monkeypox), is a zoonotic orthopoxvirus (OPV) that has historically been endemic and largely isolated to western and central Sub-Saharan Africa ^1, 2^. Other species of OPVs include cowpox (CPXV), vaccinia (VAC; smallpox vaccine), and variola virus (VAR; smallpox) which all share a high degree of genetic homology. Mpox can be transmitted through close contact with an infected person or animal, bodily fluids, respiratory secretions, or consumption of contaminated meat. Symptoms of infection can be severe and may include lymphadenopathy, fever, headache, fatigue, muscle aches, flu-like symptoms, and a characteristic rash, and death^3, 4^. In 2003 a non-endemic mpox outbreak occurred in the United States due to the importation of infected animals. In total, 71 cases of mpox were detected over 5 months. The outbreak was contained through the implementation of multiple public-health efforts including restrictions placed on the movement of exotic animals^5^. In 2022 the world faced a multi-country outbreak of non-endemic mpox, prompting the World Health Organization to announce a Public Health Emergency of International Concern. To date, over 85,000 cases with 97 deaths have been reported globally with over 30,000 of those cases and 32 deaths occurring in the United States. In contrast to the 2003 outbreak human-to-human transmission has been the primary mode of viral spread in 2022. The size and geographic distribution of the current mpox outbreak has highlighted the need for specific tools to diagnose, treat, and monitor immunity to OPVs.

OPVs are large dsDNA viruses that encode up to 200 highly conserved viral proteins resulting in extensive antigenic and serologic cross-reactivity between members of the OPV genus^6^. The protection afforded by vaccination with VAC against fatal smallpox infection is based, and dependent on the antigenic cross-reactivity between VAC and VAR. In the United States, childhood vaccination with VAC was customary until 1972, and VAC vaccination was reinstated for the US military forces in 2002 resulting in a significant number of US residents with VAC-reactive antibodies^7, 8^. More recently, JYNNEOS vaccine (Modified Vaccinia Ankara, Bavarian Nordic) has been administered to individuals deemed to be at high risk for mpox infection. Therefore, the development of a specific serologic assay to monitor mpox infection is complicated by the vaccination efforts for both smallpox and mpox^9^. The current serological assay available to detect the presence of OPV-specific antibodies is an ELISA with formalin-inactivated VAC used as the coating antigen^10-12^. The advantage of a whole-virus assay is the breadth of OPV antigens available for antibody binding – resulting in a highly sensitive assay. However, the distinct disadvantage of a whole-virus ELISA is the lack of specificity between OPV species which may give false-positive results for anyone who has received VAC vaccination. Currently, there is no approved serology assay that can discriminate between the antibodies produced following vaccination with VAC versus those mounted upon mpox infection. Therefore, we leveraged a multiplexed microsphere immunoassay (MIA) platform with the goal of maintaining OPV antigenic breadth while simultaneously gaining specificity to mpox and other non-vaccinia OPVs. In the present study, we created an algorithm using two mpox-specific peptides and 5 cross-reactive OPV antigens that detects IgG antibodies associated mpox infection with 93% sensitivity and 98% specificity. Our data support the use of serology as an important diagnostic tool for mpox infections, and as an mpox surveillance tool within vaccinated populations.

## Results

### Evaluation of Non-Vaccinia Peptides for the Specific Serologic Detection of Mpox Exposure

B21/22R is an immunomodulatory protein present in both mpox and VAR yet absent in VAC, thus allowing for serologic distinction between VAC vaccination and mpox infection^13, 14^. Dubois et al. previously reported a peptide-based ELISA using 30-mer peptides optimized for the diagnostic detection of mpox infection in populations that may include VAC vaccinated individuals. The authors reported >90% sensitivity and >90% specificity for mpox infection using 4 BSA-conjugated peptides derived from mpox (B21R-A and -B) and VAR (B22R-A and -B) in a relatively small number of mpox-positive and -negative subjects^15^. We sought to evaluate efficacy of these previously validated peptides for the diagnosis of mpox infection on our high-throughput MIA on the Luminex platform. BSA-conjugated peptides, as described in Dubois et al. were coupled to fluorescent microspheres and evaluated for IgG reactivity in the mpox-confirmed, mpox-negative, and vaccine control cohorts. Receiver Operatory Characteristic (ROC) analysis revealed moderate predictive accuracy of mpox infection for mpox-derived B21R-A and -B with an area under the curve (AUC) of 0.86 and 0.85, respectively. In contrast, VAR-derived peptides B22R-A and -B showed lower predictive accuracy for mpox infection with AUCs of 0.52 and 0.77, respectively (**Figure 1a; Supplemental Table 1**) and were eliminated from the diagnostic assay. Based on the sensitivity/specificity calculations generated by the ROC curve, we calculated clinical cutoff values for mpox peptides B21R-A and -B. Briefly, a Youden’s J index (J = sensitivity + specificity – 1) was calculated for the range of MFI values in the ROC analysis. The clinical cutoff was set as the MFI equaling the highest Youden’s J index which is equal to the best balance of specificity and sensitivity over the range of the assay (**Supplemental Table 1**).

**Figure 1:**
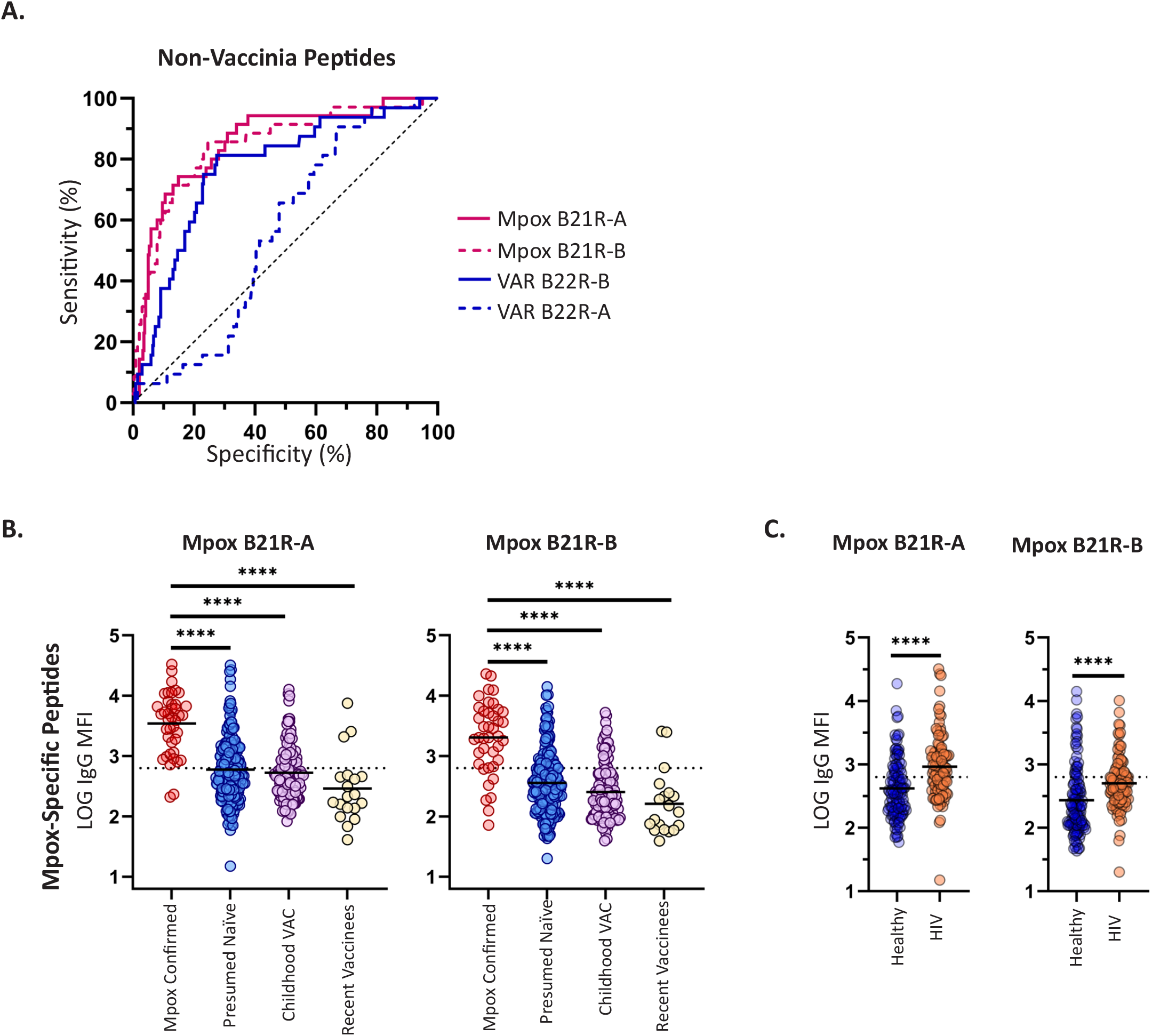
Evaluation of Non-Vaccinia Peptides for the Detection of Mpox Infection. Serum or plasma specimens from 341 presumed mpox-negative donors and 40 mpox confirmed patients were analyzed for antibody reactivity to peptide antigens derived from mpox and variola virus (VAR) by a microsphere mmunoassay. (**A)** Median fluorescence intensity (MFI) of IgG reactivity to individual 31-mer peptides was used to generate ROC curves representing for mpox-derived B21R-A and -B (magenta) or VAR-derived B22R-A and -B (blue). **(B)** The log10 MFI of IgG reactivity to mpox B21R-A and -B was plotted for mpox-negative, -positive, and vaccine control donors. The mpox-negative cohort was divided by birth year post- (presumed naive) and pre-1972 (Childhood VAC) to estimate smallpox childhood vaccination status. **(C)** Log10 MFI of IgG reactivity to mpox B21R-A and -B was plotted for mpox-negative donors post-1972 (>50) divided by “healthy” and “HIV”. Each dot represents and individual donor. Statistical significance was determined by the non-parametric Kruskal-Wallis test where *p < 0.05 **p < 0.001 ***p < 0.0001, and ****p < 0.00001 adjusted for multiple comparisons by Dunn’s test.

To visualize the baseline binding IgG reactivity to B21R-A and -B in mpox-negative donors and recent vaccinees compared to mpox confirmed donors, we plotted the log10 MFI for individual donors grouped by cohort as described in Table 1. As routine smallpox vaccination with VAC ended in the United States in 1972, consideration of birth year allowed us to estimate the childhood vaccination status of each donor in the mpox-negative cohort^8^. Therefore, mpox-negative donors were divided into “presumed naïve” and “childhood vaccination” groups based on birth year post-or pre-1972. The average MFI for the mpox-confirmed donors was significantly higher than the presumed naïve, childhood vaccination, and recent vaccinee groups for both B21R-A and -B (**Figure 1b**). However, a considerable number of mpox-negative donors show peptide-specific IgG reactivity above the clinical cutoff despite the lack of mpox exposure. Importantly, the childhood and recent vacinee groups did not show higher levels of B21R-A or -B reactivity than the presumed naïve donors indicating that smallpox vaccination does not contribute to non-specific IgG reactivity to B21R-A and -B in mpox negative populations. Since up to 57% of mpox cases reported in the 2022 outbreak have been in HIV-positive individuals^16^ we included 142 plasma or serum samples sent to the Wadsworth Center for HIV confirmatory testing in our mpox-negative cohort. HIV+ donors from the mpox-negative cohort showed a significantly higher MFI as compared to donors without HIV infection (**Figure 1c**). When considered together, the specificity of peptides B21R-A and -B (67%; **Supplemental Table 1**) is insufficient for the development of a robust mpox-specific serological assay.

**Table 1:** Cohorts Used for the Mpox Serological Assay Development.

| Cohort | Number | Naïve (#) | VAC (#) | Age (yrs) | M | F | UNK | Day |
| --- | --- | --- | --- | --- | --- | --- | --- | --- |
| **Mpox-Confirmed (Total)** | **40** | **27** | **13** | **37** | **26** | **13** | **1** | **9 - 180** |
| 2003 | 18 | 10 | 8 | 33 | 6 | 12 | 0 | 180 |
| 2022 | 22 | 17 | 5 | 40 | 20 | 1 | 1 | 9 -107 |
| **Mpox-Negative (Total)** | **341** | **221** | **121** | **40** | **170** | **170** | **1** | **n/a** |
| Healthy | 199 | 121 | 78 | 43 | 49 | 150 | 0 | n/a |
| HIV | 142 | 100 | 42 | 37 | 121 | 20 | 1 | n/a |
| **Recent Vaccinees** | **15** | **13** | **2** | **38** | **14** | **1** | **0** | **25 - 88** |
| **Specificity Panel** | **115** | **-** | **-** | **-** | **-** | **-** | **115** | **n/a** |

**Table 2:**
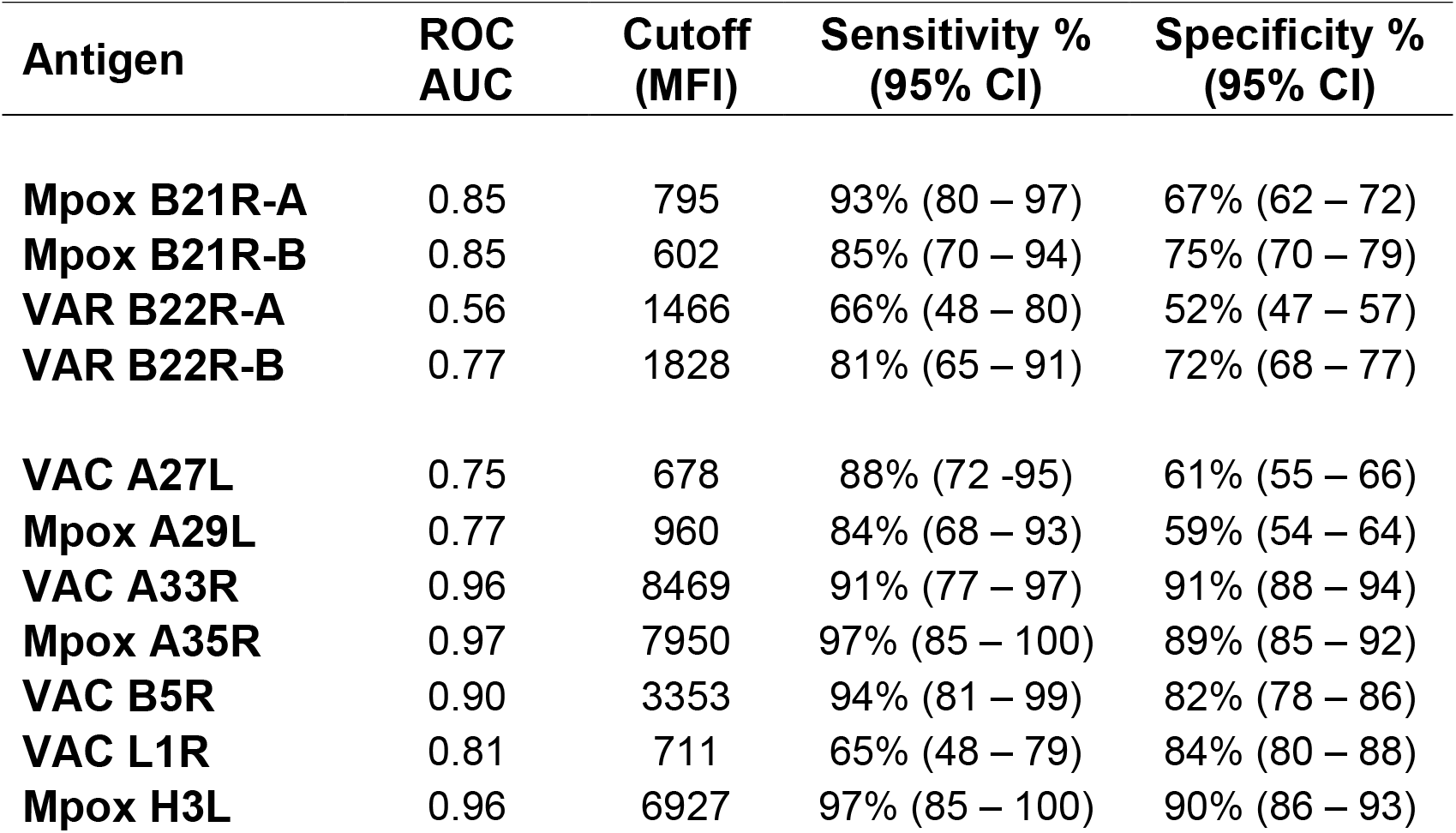
Diagnostic Performance of OPV Peptides and Antigens.

| Antigen | ROC AUC | Cutoff (MFI) | Sensitivity % (95% CI) | Specificity % (95% CI) |
| --- | --- | --- | --- | --- |
| **Mpox B21R-A** | 0.85 | 795 | 93% (80 – 97) | 67% (62 – 72) |
| **Mpox B21R-B** | 0.85 | 602 | 85% (70 – 94) | 75% (70 – 79) |
| **VAR B22R-A** | 0.56 | 1466 | 66% (48 – 80) | 52% (47 – 57) |
| **VAR B22R-B** | 0.77 | 1828 | 81% (65 – 91) | 72% (68 – 77) |
| **VAC A27L** | 0.75 | 678 | 88% (72 -95) | 61% (55 – 66) |
| **Mpox A29L** | 0.77 | 960 | 84% (68 – 93) | 59% (54 – 64) |
| **VAC A33R** | 0.96 | 8469 | 91% (77 – 97) | 91% (88 – 94) |
| **Mpox A35R** | 0.97 | 7950 | 97% (85 – 100) | 89% (85 – 92) |
| **VAC B5R** | 0.90 | 3353 | 94% (81 – 99) | 82% (78 – 86) |
| **VAC L1R** | 0.81 | 711 | 65% (48 – 79) | 84% (80 – 88) |
| **Mpox H3L** | 0.96 | 6927 | 97% (85 – 100) | 90% (86 – 93) |

### Evaluation of OPV Antigens for the Serologic Detection of Mpox Exposure

We asked whether any naturally cross-reactive recombinant proteins derived from VAC or mpox could aid in the development of an mpox-specific serologic assay. The predictive accuracy of different OPV antigens for the detection of mpox infection was tested with a variety of VAC and mpox-derived protein antigens that were available through a public repository, in-house production, or commercial vendors. Recombinant proteins VAC A27L, mpox A29L, VAC A33R, mpox A35R, VAC B5R, VAC L1R, and mpox H3L were tested for IgG antibody reactivity using the same mpox-infection confirmed and mpox-negative samples as described in Figure 1 (**Figure 2a**). ROC analysis of each antigen revealed distinct differences in the predictive accuracy for mpox infection. VAC A33R, mpox A35R, VAC B5R, VAC L1R and mpox H3L showed moderate to excellent predictive accuracy with AUC values above 0.80^17^. In contrast, VAC A27L and homologous mpox A29L showed low predictive accuracy with AUCs below 0.80 and were removed from the assay. As with the mpox-specific peptides, clinical cutoffs were calculated for the remaining antigens using the Youden’s J index calculated from the ROC analysis.

**Figure 2:**
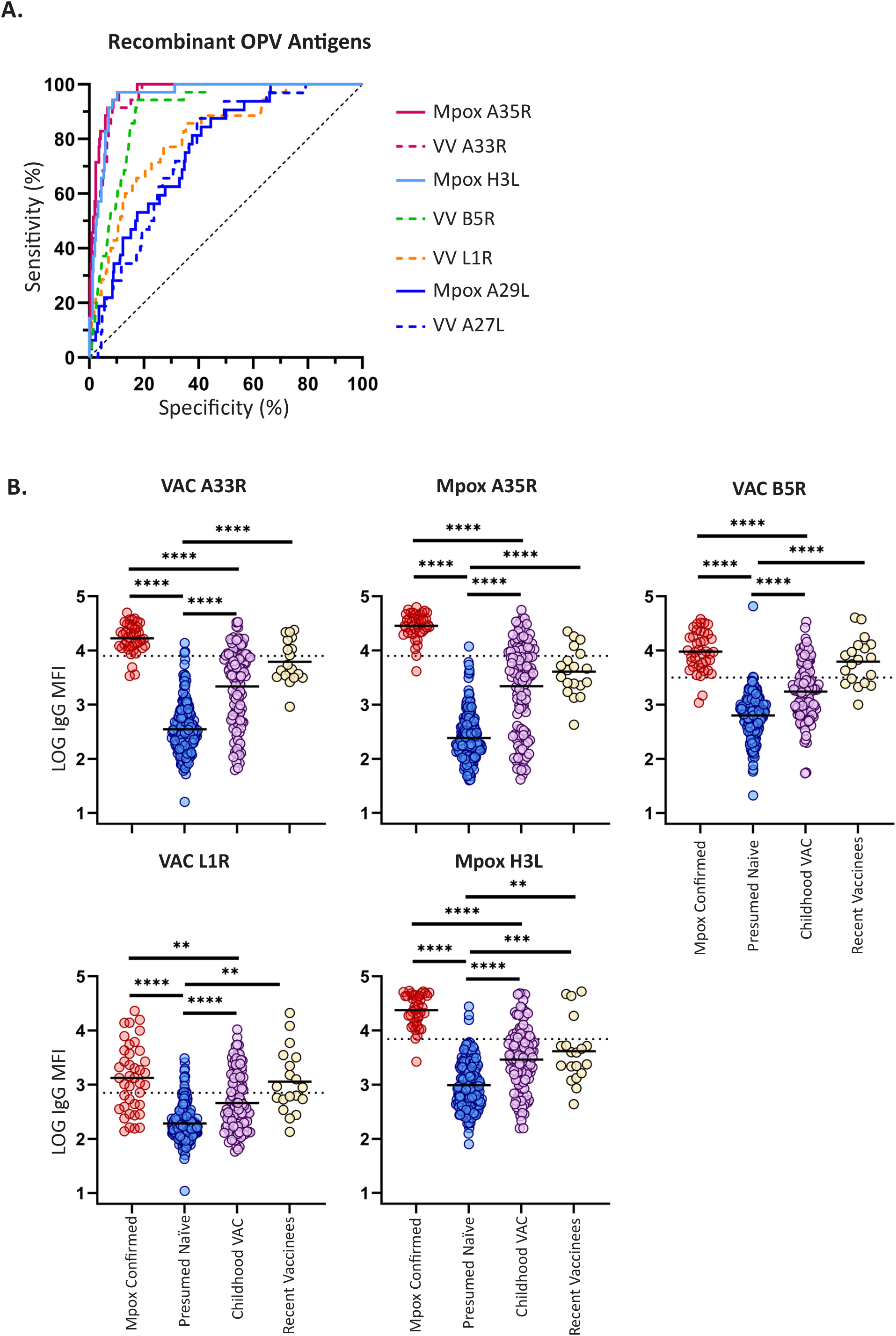
Evaluation of Orthopoxvirus Antigens for the Detection of Mpox Infection. Serum or plasma specimens from 341 presumed mpox-negative donors and 40 mpox confirmed patients were analyzed for antibody reactivity to recombinant protein antigens derived from mpox or vaccinia virus (VAC) by microsphere immunoassay. **(A)** Median fluorescence intensity (MFI) of IgG reactivity to recombinant antigens from mpox or VAC (mpox A35R, solid magenta; VAC A33R, dashed magenta; mpox H3L solid light blue; VAC B5R dashed green; VAC L1R dashed orange; mpox A29L solid dark blue, VAC A27L dashed dark blue) were used to generate ROC curves used to represent the sensitivity (%) and specificity (%) of each antigen to detect mpox infection. (B) Log10 MFI of IgG reactivity to OPV antigens (A33/35, B5R, L1R, and H3L) plotted for mpox-negative, -positive, and recent vaccinee donors. Each dot represents and individual donor. Statistical significance was determined by the non-parametric Kruskal-Wallis test where *p < 0.05 **p < 0.001 ***p < 0.0001, and ****p < 0.00001 adjusted for multiple comparisons by Dunn’s test.

To visualize the baseline binding IgG reactivity to VAC A33R, mpox A35R, VAC B5R, VAC L1R, and mpox H3L in mpox-negative donors and recent vaccinees as compared to mpox confirmed donors, we plotted the log10 MFI for individual donors grouped by cohort as described in Figure 1. The average MFI of the mpox-confirmed donors for each recombinant antigen was significantly higher than the MFI’s for the mpox-negative presumed naïve and childhood vaccination groups. As expected, the childhood vaccination group had significantly higher average MFI’s than the presumed naïve group indicating previous exposure to VAC antigens and durable humoral memory from childhood smallpox vaccination ^14, 18^ (**Figure 2b)**. Although not statistically significant, the average MFI of recent vaccinees trended higher than the childhood vaccination group across all recombinant antigens. These data show that mpox infection can induce a more robust antibody response than vaccinia vaccination, which may be exploited for the development of a diagnostic serology assay.

### Distinct Pattern of IgG Reactivity to OPV Antigens and Mpox-specific Peptides Between Mpox-confirmed and Mpox-negative Donors

We asked whether multiplexing multiple antigens would reveal patterns of serological reactivity that may be used to distinguish between VAC vaccination and mpox infection. To this end, we plotted each specimen/antigen combination on a heat map to visualize the pattern of reactivity in the mpox-confirmed and mpox-negative cohorts. The mpox-confirmed donors showed uniform and simultaneous reactivity to the mpox peptides and the OPV antigens that were tested (**Figure 3**). In contrast, the mpox-negative presumed naïve group showed sporadic reactivity to the mpox peptides and OPV antigens. Here, individuals that showed positive reactivity to the B21R peptides did not show simultaneous reactivity with the OPV antigens. The mpox-negative childhood vaccination group displayed an increased frequency of antigen reactivity as expected with previous exposure to VAC, yet rarely showed simultaneous reactivity to the mpox peptides and OPV antigens. Recent vaccinees displayed a similar pattern of mpox peptide and OPV antigen reactivity to the childhood vaccination group. The differential patterns of antigen reactivity between mpox-confirmed and mpox-negative cohorts suggest a combination of OPV antigens and mpox-specific peptides may provide specific serologic detection of mpox infection.

**Figure 3:**
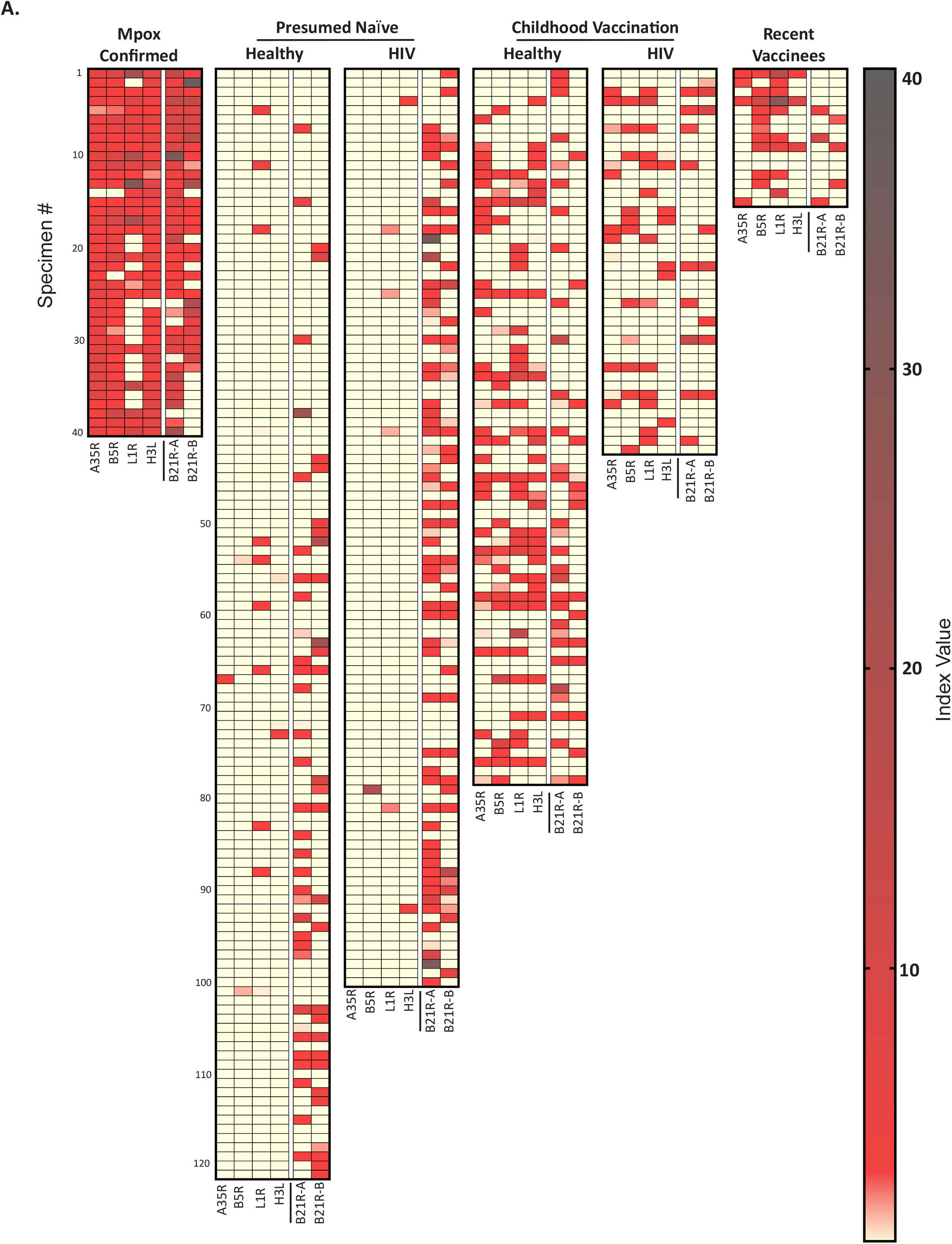
Distribution of OPV Antigen and Mpox Peptide Reactivity Among Negative and Mpox Cohorts. Heat maps indicate the relative index value (MFI/cutoff value) of each antigen (Mpox A35R, VV B5R, VV L1R, Mpox H3L, Mpox B21R-A, and Mpox B21R-B; vertical columns) for each serum/plasma donor (horizontal rows). Yellow corresponds to a negative index value (< 1.0) below the clinical cutoff. The degree of red saturation corresponds to higher index values (> 1.0) above the clinical cutoff.

### Development of an Algorithm for the Specific Detection of Mpox Infection

With the goal of developing an algorithm for a mpox-specific assay we counted the number of clinically reactive (MFI above the clinical cutoff) OPV antigens for every individual in the mpox-confirmed and -negative cohorts. Donors from the mpox-confirmed cohort were simultaneously reactive for at least 3 OPV antigens, with a median count of 4 antigens (**Figure 4a**). In contrast, presumed naïve donors from the mpox-negative cohort were rarely reactive for 1 OPV antigen with a median count of 0 antigens. Mpox-negative donors from the childhood vaccination and recent vaccinee groups showed a range of reactivity from 0 to 4 OPV antigens with median counts of 0 and 2.0, respectively. Setting a clinical cutoff of 3 or more reactive antigens captures 98% of the mpox-confirmed donors while excluding 95% of the mpox-negative cohort.

**Figure 4:**
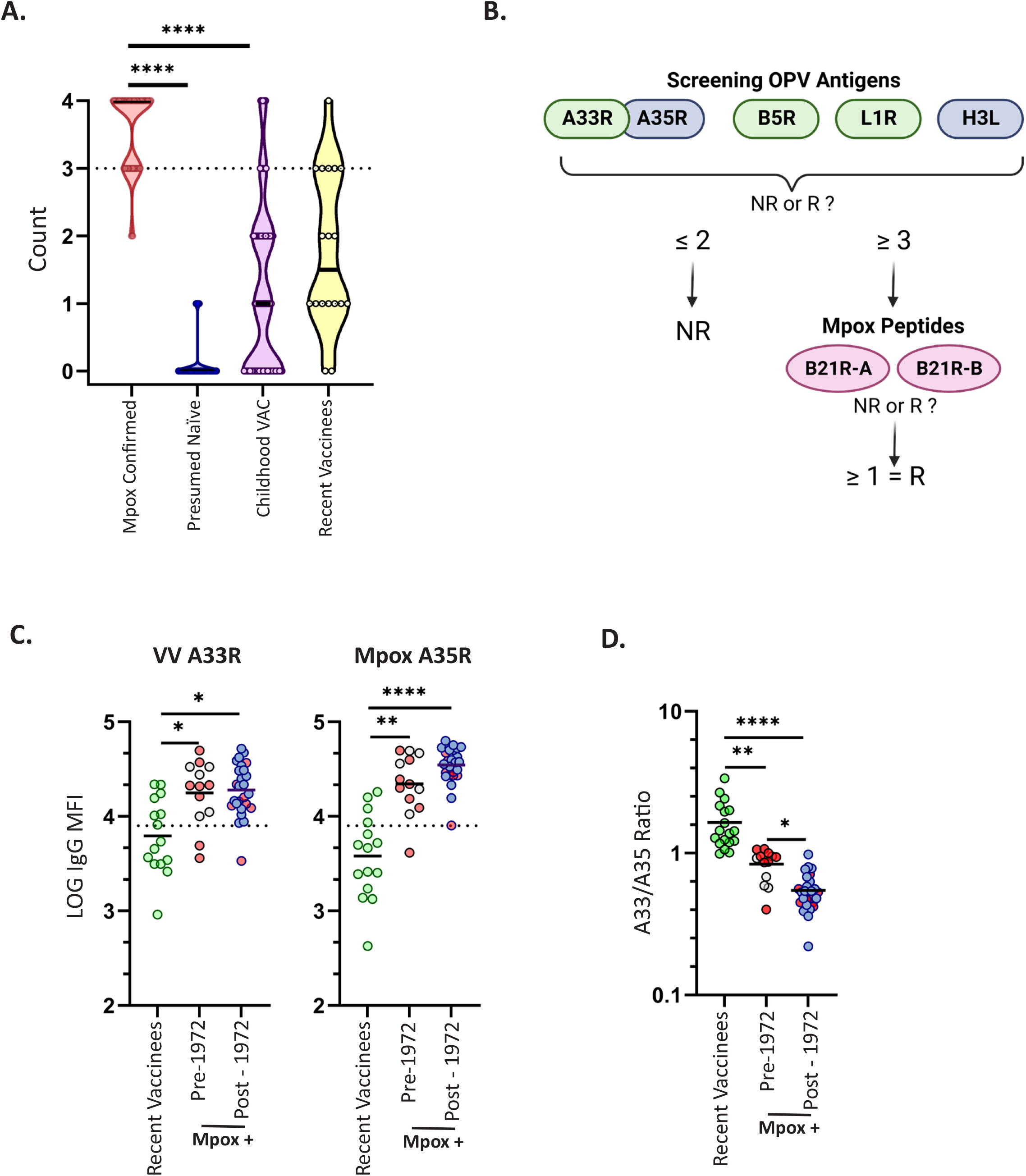
OPV Antigen Count and Algorithm for the New York State Orthopoxvirus (non-vaccinia) Microsphere Immunoassay for IgG Antibody Detection **(A)** The number of OPV antigens (A33/35, B5R, L1R, and H3L) counted as reactive (R). Reactivity was defined as an MFI value at or above the calculated clinical cutoff value. The antigen count distribution was displayed as a violin plot for mpox-confirmed and mpox-negative cohorts. The mpox-negative cohort divided by birth year post- (presumed naïve) and pre-1972 (Childhood VAC) to estimate small-pox childhood vaccination status. **(B)** Serum or plasma specimens tested for mpox infection using a multiplexed microsphere immunoassay will be subjected to a 2-tier algorithm using a combination of mpox-specific peptides and OPV antigens. First, specimens will be considered for IgG reactivity to a set of cross-reactive OPV antigens (VAC A33R, mpox A35R, VAC B5R, VAC L1R, and mpox H3L). Samples with reactivity to 2 or fewer antigens will be considered non-reactive (NR; MFI < clinical cutoff) overall for mpox exposure while samples that test reactive (R; MFI > clinical cutoff) to 3 or more OPV antigens will move to the second tier of the algorithm. Next, samples will be considered for their reactivity to mpox peptides B21R-A and -B. Samples with no peptide reactivity will be considered NR for mpox infection. Samples with reactivity to one or more mpox peptides will be considered R for mpox infection. **(C)** Serum specimens from mpox-confirmed donors (n=40) and recent vaccinees (n=15) were analzyed for IgG reactivity to viral homologues VAC A33R and mpox A35R. Mpox confirmed donors were separated by birth year to simulate vaccination status before infection. Green circles indicate recent vaccinees. Grey circles indicate a birth year prior to 1972 (childhood vaccine), and blue circles indicate a birth year after 1972 (unvaccinated). Red circles in both groups indicate individuals infected in the 2003 mpox outbreak. The dashed line in each plot indicates the reactivity cutoff for the indicated antigen as determined by ROC curve. **(D)** Serum IgG reactivity ratio representing the mpox A35R MFI divided by the VAC A33R MFI. Green circles indicate recent vaccinees. Grey circles indicate a birth year prior to 1972 (childhood vaccine), and blue circles indicate a birth year after 1972 (unvaccinated). Red circles in both groups indicate individuals infected in the 2003 mpox outbreak. Statistical significance was determined by the non-parametric Kruskal-Wallis test where *p < 0.05 **p < 0.001 ***p < 0.0001, and ****p < 0.00001 adjusted for multiple comparisons by Dunn’s test.

Based on the high antibody reactivity of mpox-infected donors to both cross reactive OPV antigens and the mpox-specific peptides, we developed a two-tiered algorithm designed to remove individuals without true mpox infection from further analysis (**Figure 4b**). First, the total number of reactive (MFI ≥ cutoff) OPV antigens (A33/35R, B5R, L1R, H3L) was counted for each individual donor. Donors with less than 3 reactive OPV antigens will automatically be considered negative (non-reactive; NR) for mpox-infection. Donors with 3 or more reactive OPV antigens will be considered for the next step of the algorithm. At this stage 95% of the mpox-negative donors (n=341) have been labeled NR and removed from the analysis, while 98% of the mpox-infected cohort will move to the next tier of the algorithm. Next, donors that “passed” the first step of the algorithm will be considered for IgG reactivity to the mpox-specific peptides B21R-A and -B. Finally, donor samples that are reactive (MFI ≥ cutoff) for one or both peptides will be considered positive (reactive; R) for mpox infection. After the second step of the algorithm 37 out of 40 patients from the mpox-confirmed cohort have been labelled R, giving our diagnostic test (New York State Orthopoxvirus Non-Vaccinia Virus Microsphere Immunoassay for IgG Antibody Detection; NYS-OPV-MIA) 93% (95%CI = 80-97%) sensitivity for mpox infection with an overall specificity of 98% (95%CI = 96-99%) (**Table 3**). It is important to note that assay specificity is different depending on the test population. When testing vaccinated donors assay specificity is 94% (95%CI = 88-97%) for the childhood vaccinee group, and 93% (95%CI = 88-96%) when the recent vaccinees are added to the calculation. However, the NYS-OPV-MIA is highly (100%; 98-100%) specific when testing unvaccinated individuals under 50 years of age. As part of assay validation we also tested a specificity panel of 115 serum specimens with known antibodies to a diverse group of bacterial and viral pathogens, as well as anti-nuclear antibodies (ANA) and rheumatoid factor (RF) (Supplemental Table 1). The specificity panel included specimens from infections that may be confused with mpox, including Syphilis (n=8), Herpes Simplex Virus (HSV; n = 15), and Varicella Zoster (n = 18). When combined with the mpox-negative cohort (n = 456) the specificity of the NYS-OPV-MIA remains at 98% (95%CI = 96-99%). In summary, we have developed a robust serological assay with high sensitivity and high specificity to aid in the diagnosis of mpox-infection.

**Table 3:**
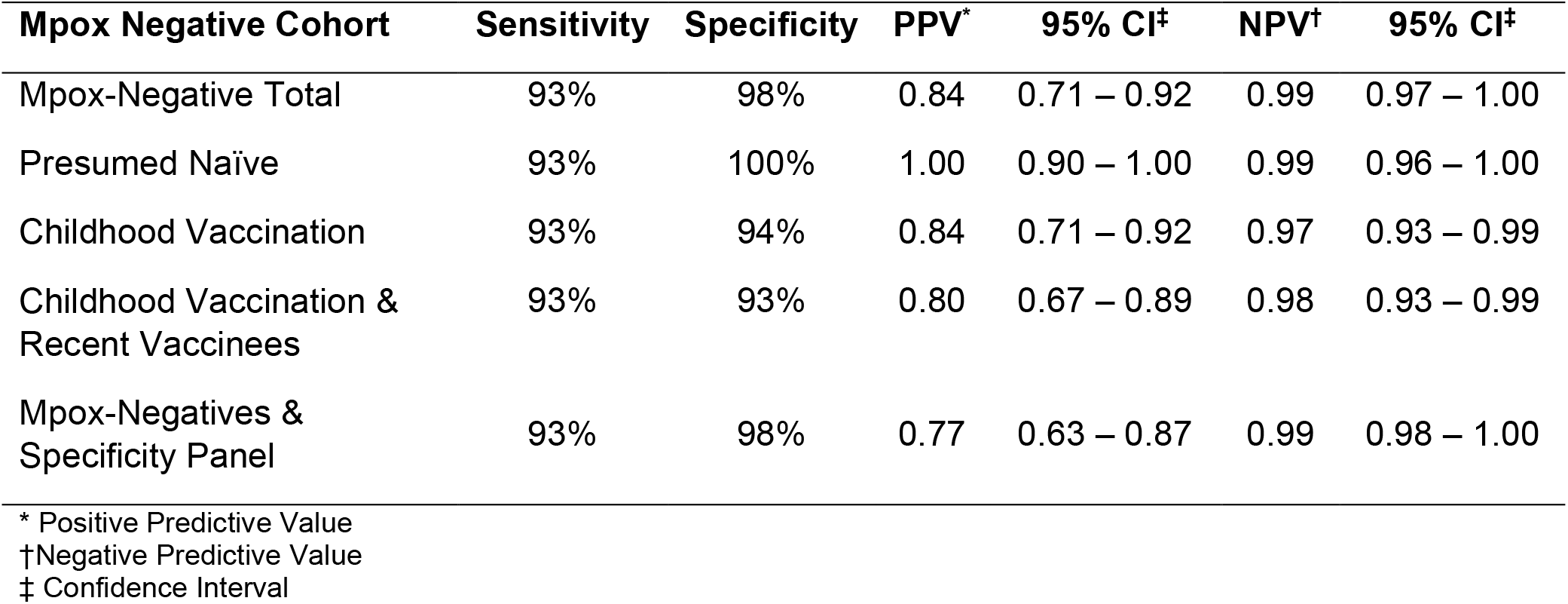
Sensitivity and Specificity of the NYS-OPV-MIA Based on Age Group.

| **Mpox Negative Cohort** | **Sensitivity** | **Specificity** | **PPV\*** | **95% CI‡** | **NPV†** | **95% CI‡** |
| --- | --- | --- | --- | --- | --- | --- |
| Mpox-Negative Total | 93% | 98% | 0.84 | 0.71 – 0.92 | 0.99 | 0.97 – 1.00 |
| Presumed Naïve | 93% | 100% | 1.00 | 0.90 – 1.00 | 0.99 | 0.96 – 1.00 |
| Childhood Vaccination | 93% | 94% | 0.84 | 0.71 – 0.92 | 0.97 | 0.93 – 0.99 |
| Childhood Vaccination &<br>Recent Vaccinees | 93% | 93% | 0.80 | 0.67 – 0.89 | 0.98 | 0.93 – 0.99 |
| Mpox-Negatives &<br>Specificity Panel | 93% | 98% | 0.77 | 0.63 – 0.87 | 0.99 | 0.98 – 1.00 |

\* Positive Predictive Value † Negative Predictive Value ‡ Confidence Interval

### Strength of Mpox Antigen Binding is Dictated by the First Viral Exposure

The induction of potent cross-protective immunity through vaccinia virus vaccination for protection against smallpox is well established^19, 20^. There is also strong evidence for VAC vaccination to provide some degree of protection against mpox infection^21, 22^. Our mpox serology assay includes the homologous proteins A33/35R from both VAC and mpox respectively, which allowed us to ask how previous exposure to VAC through vaccination influences antibody binding following mpox infection. To this end, we divided the mpox cohort by birth year to reflect the childhood vaccination status of the mpox-positive cohort. When considering IgG binding to VAC A33R, mpox-infected patients bind equally well to VAC A33R regardless of previous VAC exposure. In contrast, mpox-infected patients under 50 trended toward higher binding to mpox A35R as compared to their previously vaccinated counterparts (**Figure 4c**). To estimate the relative binding of VAC A33R versus mpox A35R in each antigen exposure group we calculated a binding ratio by dividing the IgG MFI of VAC A33R by the IgG MFI of mpox A35R. The resulting data revealed differential binding of the A33/35R homologues based on the primary immunizing virus. The vaccine control group showed the highest VAC/mpox binding ratio indicating that antibodies from this group bound the VAC A33R protein better than the mpox homologue. In contrast, previously unvaccinated mpox-infected patients showed the lowest binding ratio, indicating better antibody binding to mpox A35R rather than the VAC homologue. Interestingly, previously vaccinated mpox-infected patients showed an intermediate A33/35R binding ratio suggesting a contribution from both VAC-derived and mpox-derived antibodies to the overall binding profile in these patients. Together these data demonstrate the presence of distinct epitopes in homologous proteins despite high levels of cross-reactivity, and that original antigenic sin may play a role in the humoral response to mpox in previously vaccinated individuals.

## Discussion

In response to a Public Health Emergency of International Concern, we developed a sensitive (93%) and specific (98%) multiplex MIA for the differential serologic detection of mpox infection in VAC vaccinated populations. The assay contains 2 mpox-specific peptides and 5 cross-reactive OPV recombinant proteins derived from either VAC or mpox. Here, we leveraged both multiplex technology and the differential antibody reactivity profiles resulting from VAC vaccination and mpox infection to build specificity into our serologic assay.

In 2012 Slifka and colleagues reported a highly sensitive mpox-specific ELISA based on 30-mer peptides derived from the B21/22R gene that is present in mpox yet absent in VAC^15^. We tested the BSA-conjugated B21/22R peptides on our multiplex MIA as a high-throughput alternative to the previously described ELISA. Of the four peptides tested, mpox-derived B21R-A and -B had the best discriminatory capacity for mpox infection, where VAR-derived B22R-A and -B were comparatively poor. In contrast to the Dubois-Slifka study, we found extensive non-specific IgG reactivity to the peptides in an mpox-negative population, the origin of which is unknown. The most likely explanation for the discrepant specificity between studies is the size and composition of our negative cohort. The mpox-negative cohort of the Dubois-Slifka study was composed of 20 healthy naïve, recently vaccinated, and childhood vaccinated donors for a total of 60 test subjects. Here, we calculated diagnostic specificity of the B21/22R peptides based on a negative cohort of 341 donors, 142 of which were HIV-positive. A thorough analysis of our mpox-negative cohort revealed that much of the non-specific reactivity seen to the peptides was present in the samples collected from people living with HIV. In fact, non-specific serologic reactivity in HIV+ populations due to polyclonal B cell activation and hypergammaglobulinemia is a well-characterized aspect of HIV infection^23, 24^. Further, many HIV-1-specific broadly neutralizing antibodies generated during HIV infection are polyreactive^25^. The high frequency of HIV+ patients in the mpox test population^16^ has the potential to impact assay performance and underscores the importance of considering test population demographics prior to assay development. In addition, we clearly demonstrated that the B21/22R-derived peptides tested here had insufficient specificity to be used as a stand-alone diagnostic assay.

With a lack of additional non-vaccinia derived proteins to test, we turned to immunodominant VAC and mpox antigens known for their cross-reactivity across OPV species. We demonstrate that mpox-infected donors have robust IgG reactivity to the immunodominant antigens A33/35R, B5R, L1R, and H3L. In contrast, both recent and childhood vaccinees show detectable antibody reactivity to one or more of the antigens tested, while the antibody profile of each individual lacks the magnitude and antigenic breadth that is seen following mpox infection. Our data is supported by previous work showing the majority of VAC vaccinees recognize a small fraction of the OPV proteins tested in a protein microarray. Moreover, no single antigen in the array was universally recognized by all vaccinees^12^. Further, antibodies specific for immunodominant antigens A33, B5, L1, and H3 are rarely seen in all individuals following primary VAC immunization^12, 26, 27^. The cause for reduced antibody titer and antigenic breadth in vaccinated versus mpox infected individuals may be due to differences in viral load or tropism that occur through vaccination versus infection^28^. In fact, challenge models have shown increases in both the magnitude and coverage of OPV-specific antibodies following mpox infection of VAC vaccinated animals^12^.

Serologic assays measure the host response to an infectious agent which can be detected for months to years following infection. However, measuring the host immune response comes with inherent challenges such as antigenic cross-reactivity and genetic variability between individuals. We demonstrated that a multiplex immunoassay platform using a combination of mpox peptides and OPV proteins can identify mpox infection with high sensitivity and specificity in a population where VAC vaccination has occurred. Furthermore, we show that mpox infection and VAC vaccination result in differential patterns of IgG reactivity that could be exploited for diagnostic detection despite high levels of antigenic cross-reactivity. The bead-based platform described in this study is amenable to high-throughput processing which may be useful for performing large-scale sero-epidemiology studies^29^ in high-risk populations and inform our understanding of mpox transmission in the future.

### Limitations of the Study

The B21/22R gene is present used as the final discriminator for mpox infection in our assay is expressed in other non-vaccinia OPV species. Interpretation of our assay may be difficult in regions where cowpox or other non-vaccinia OPVs are endemic^30^. In addition, sequencing efforts during the 2002 mpox outbreak have identified isolates with genetic deletions that may affect the B21R gene^31^. B21R deletions in the mpox genome have the potential to effect assay sensitivity if such mutations were to be commonplace in circulating mpox strains.

## Data Availability

All data produced in the present study are available upon reasonable request or contained in the present manuscript.

## Acknowledgements

We wish to acknowledge and express our gratitude to all of the study participants for their generosity, without wich this study would not be possible. We acknowledge and thank the following members of the Diagnostic Immunology Laboratory for their expert technical assistance: Kelly Howard, Kyle Carson, and Theresa Lamson. We acknowledge and thank Jean Rock and Rachel Bievenue of the Bloodborne Viruses Laboratory and Michael Perry of the Biodefense Laboratory for providing sample preparation. We acknowledge and thank Heidi Dillenbeck for quality control and quality assurance support. We thank Archana Thomas of the Slifka Laboratory for assistance with sample preparation and shipping. We thank Dr. Charles Gonzalez of the AIDS Institute, NYSDOH for facilitating serum specimen acquisition. We thank Nicholas J. Mantis for critical reading of the manuscript. Peptides were manufactured and supplied by Aalto Bio Reagents, Biosynth Group.

## Author Contributions Statement

**Conceptualization**, J.L.Y., W.T.L., M.K.S.; **Methodology**, D.T.H., J.L.Y.; **Validation**, D.T.H., K.E.K.; **Formal Analysis**, J.L.Y. **Investigation**, D.T.H., J.L.Y.; **Resources**, K.C., L.S., S.T.C., G.Y.C., M.C.B-G., G.K., D.A., K.S., PVI, V.S., D.F., J.M., W.H., P.N., C.E., M.K.S.; **Data Curation**, J.L.Y., D.T.H. **Writing – Original Draft**, J.L.Y.; Writing – Review and Editing, J.L.Y, M.K.S., W.T.L and N.J.M.; **Visualization**, J.L.Y.; **Supervision**, W.T.L, M.K.S., and J.L.Y.

## Materials and Methods

### Experimental Model and Subject Details

#### Human Subjects

This assay development study was performed using deidentified sera and plasma for a public health function in a declared Public Health Emergency (PHE). It has been deemed “Non-human subject research” by the NYS Institutional Review Board.

Biospecimens provided by the Icahn School of Medicine at Mount Sinai from the Personalized Virology Initiative were reviewed and approved by the Mount Sinai Hospital Institutional Review Board (IRB-16-16772 and IRB-16-00791). All participants provided written informed consent before specimens and clinical information were collected. All specimens were coded before processing and analysis.

#### Specimen Collection

Whole blood in a red top/SST or equivalent tube, specimens allowed to clot at room temperature, centrifuged and transferred serum into plastic tube(s), with storage refrigerated at 2-8°C prior to shipment on freezer

#### Mpox-Negative Cohort

##### Healthy-Normal

Wadsworth Center employees volunteered serum specimens for assay development (n=114); 59 were born prior to 1972 and 55 born after 1972. Normal Healthy Serum Pre-COVID-19 Pandemic Panel E was purchased from Access Biologicals (n=85); 85 were born prior to 1972 and 66 born after 1972 with military status unknown.

#### HIV+

One hundred and fourty-three (142) remnant plasma and serum specimens submitted for HIV diagnostic testing to the Wadsworth Center between 1/1/2018 and 12/31/2021. Specimens were submitted for a variety of reasons, including confirmation of a reactive result on an HIV rapid test, to resolve discordant HIV test results, or to verify a previous HIV diagnosis. HIV status was ‘HIV-1 antibody positive for 131 (92%) and ‘HIV antibody negative for 12 (8%) of the specimens. Specimens were collected from individuals born either prior to 1972 (n=42) or after 1980 (n=100).

#### Mpox-Confirmed Cohort

The mpox-confirmed cohort (n = 40) is composed of specimens collected during the 2003 and 2022 United States outbreaks (**Table 1; Supplemental Figure 1**).

##### 2003 Outbrea

Eighteen (18) of the 40 samples were collected following the 2003 mpox outbreak in the United States at approximately 6 months post-infection as previously described^21^. Eight out of 18 donors (44%) received childhood smallpox immunization. The 2003 cohort was 66% female with an average age of 33 years at serum collection^21^.

##### 2022 Outbreak

Twenty-two (22) of the 40 samples were collected during the 2022 mpox outbreak. In total, the group was 90% male with an average age of 40 years at specimen collection.

Two (2) serum specimens were submitted to the Wadsworth Center Biodefense Laboratory for mpox diagnostic testing on days 9 and 15 post-symptom onset, respectively. Mpox diagnosis was confirmed by PCR assay.

At the Zucker School of Medicine at Hofstra/Northwell, serum was collected from 13 persons with confirmed mpox infection, as defined by a positive PCR assay result. Specimens were de-identified and the data collected included age, gender, date of mpox symptom onset, date of PCR positive test, name of PCR assay used, VAC vaccination history, and date of serum collection.

At the Icahn School of Medicine at Mt. Sinai, serum was collected from 7 persons with confirmed mpox infection by PCR assay. Specimens were de-identified and the shared data included age range, gender, days since symptom onset, and VAC vaccination history.

#### Recent Vaccinee Cohort

The vaccinee cohort is composed of 5 serum specimens from Wadsworth Center employee donors who were vaccinated with JYNNEOS® (day 56 post-primary inoculation), and 10 serum specimens provided by donors from The Ican School of Medicine at Mt. Sinai that were vaccinated with JYNNEOS® (n=8) or ACAM 2000 (n=2). All serum specimens used for analysis were collected at least 20 days following the last dose of vaccine.

### Expression, Production, and Purification of Antigens

#### Production of B21/22R Peptides

Peptides were manufactured and supplied by Aalto Bio Reagents, Biosynth Group. The sequences for peptides MPV B21R-A and -B, and VM B22R-A and B, were described in Dubois et. al.^15^ MPV B21R-A and B correspond to B21R.179/180 and B21R.185/186, respectively. VM B22R-A and B correspond to B22R.64/65 and B22R.82/83, respectively. Peptide produced using standard Fmoc-based solid phase synthesis and purified using reverse phase high-performance liquid chromatography (RP-HPLC) to >90% pure as determined by HPLC and/or LC/MS. Conjugation to BSA occurred through the side chain of cysteine via sylfhydryl chemistry by mixing peptides with maleimide-activated BSA for two hours at room temperature.

#### Recombinant OPV Antigens

Recombinant proteins were obtained from several commercial sources. Recombinant A27L (VAC-WR-A27L; NR 2213), A33R (VAC-WR-A33R; NR545), B5R (VAC-WR-B5R; NR-546) and L1R (VAC-WR-L2R; NR-21986) were obtained from BEI Resources, Manassas VA. Recombinant mpox A35R (230-30238) and mpox H3L (230-30233) were purchased from Ray Biotech, Peachtree Corners, GA. Mpox A29L was produced by the Wadsworth Center Protein Expression Core. The mpox A29L gene (GenBank: AY160186) subcloned into the plasmid pETMPOX/A27Lo-His6 was obtained from bei Resources (Catalog # NR-3022). The construct was transformed into BL21(DE3)pLysS (Novagen) and expressed in Luria Broth containing 30 μg/mL kanamycin. Cell pellets were lysed in 50 mM Tris HCl, 300 mM NaCl, 10 mM imidazole, 0.1% Triton X100, 10% sucrose pH 7.5 containing EDTA-free Complete Protease inhibitor (Roche) and 50 U Benzonase (Novagen). Cleared lysate was applied to a 1 ml HisTrap HP column (Cytiva) in 50 mM Tris HCl, 150 mM NaCl, 10% glycerol pH 7.5, washed with 50 mM Tris HCl, 1 M NaCl, 0.25 M arginine pH 8.5, and eluted with a linear gradient over 20 column volumes to 50 mM Tris HCl, 150 mM NaCl, 10% glycerol, 500 mM imidazole pH 7.5. Fractions containing protein were pooled and dialyzed against 50 mM Tris HCl, 150 mM NaCl, 10% glycerol, 1 mM EDTA pH 7.5 and then twice against a large excess of phosphate-buffered saline (PBS). The protein was checked for size (16.4 kDa) and purity by sodium dodecyl sulfate polyacrylamide electrophoresis (SDS-PAGE).

#### OPV-specific Multiplex Microsphere Immunoassay (MIA)

Specimens were assessed for the presence of antibodies reactive to OPV antigens using an MIA. Recombinant proteins were covalently linked to the surface of fluorescent, magnetic microspheres (Luminex Corporation). Serum or plasma samples (25 μl at 1:100 dilution) and antigen-coupled microspheres (25 μl at 5×10^4^ microspheres/mL) were mixed and incubated for 30 minutes at 37°C. Serum-bound microspheres were washed and incubated with phycoerythrin (PE)-conjugated secondary antibody specific for human IgG (Southern Biotech). After washing and final resuspension in buffer, the samples were analyzed on a FlexMap 3D analyzer using xPONENT software, version 4.3 (Luminex Corporation).

**Supplemental Table 1:** Specificity Panel for Cross-Reactivity Testing of the NYS-OPV-MIA.

| Antibody Positive Sera | Number Tested | Positive (R) | Negative (NR) |
| --- | --- | --- | --- |
| Anti-nuclear antibody (ANA) | 5 | 0 | 5 |
| *Anaplasma phagocytophilum* | 3 | 1 | 2 |
| *Bartonella spp.* | 1 | 0 | 1 |
| *Borrelia burgdorferi* | 7 | 0 | 7 |
| *Brucella spp.* | 3 | 0 | 3 |
| California Serogroup Viruses | 4 | 0 | 4 |
| Cytomegalovirus | 5 | 1 | 4 |
| *Coxiella burnetii* | 3 | 0 | 3 |
| Dengue Virus | 3 | 0 | 3 |
| Epstein-Barr Virus (EBV) | 3 | 0 | 0 |
| *Ehrlichia chaffeensis* | 3 | 0 | 3 |
| Enterovirus | 2 | 0 | 2 |
| Hepatitis B Virus | 1 | 1 | 0 |
| Hepatitis C Virus | 2 | 0 | 2 |
| HIV1 | 2 | 0 | 2 |
| Herpes Simples Virus | 13 | 0 | 13 |
| Measles | 5 | 0 | 5 |
| Mumps | 3 | 0 | 3 |
| Varicella-Zoster | 18 | 0 | 18 |
| Parvovirus | 3 | 0 | 3 |
| Powassan Virus | 3 | 0 | 3 |
| Rheumatoid factor (RF) | 2 | 0 | 2 |
| *Rickettsia spp.* | 2 | 0 | 2 |
| Rubella | 2 | 0 | 2 |
| Syphilis | 8 | 0 | 8 |
| Western/Eastern Equine Encephalitis virus | 1 | 0 | 1 |
| West Nile Virus | 6 | 1 | 5 |
| Yellow Fever Virus | 1 | 0 | 1 |
| Zika Virus | 2 | 0 | 2 |

**Supplemental Figure 1:**
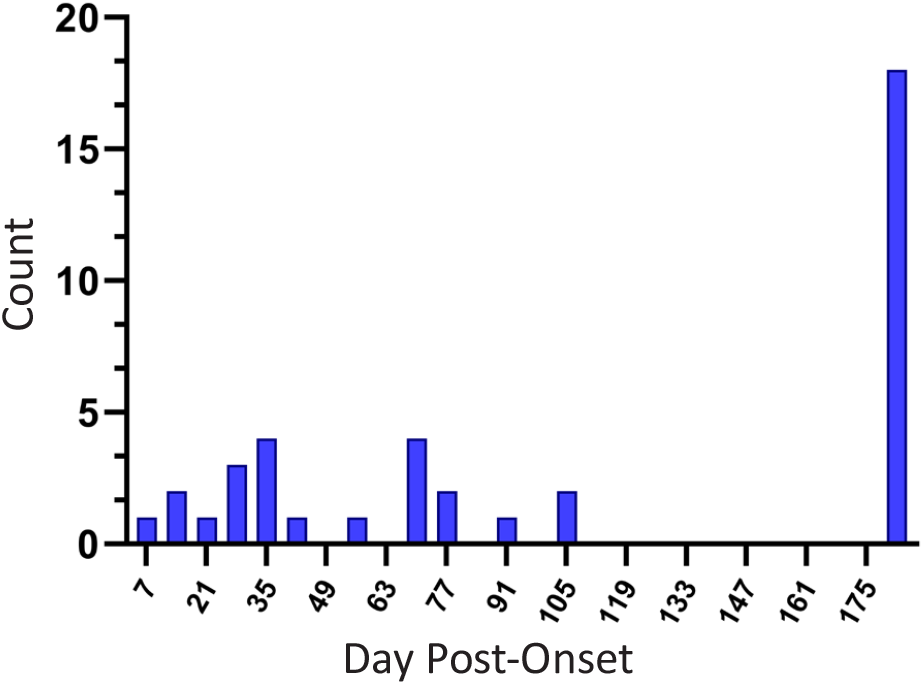
Distribution of Days Post-Onset for Mpox-Confirmed Cohort. Histogram represents the number of specimens/donors at each day post-onset for the mpox-confirmed ohort. The 18 specimens at 180 days post-onset are from the 2003 group and the remainder of specimens re from the 2022 outbreak.

## Main Text References

1. Rimoin AW, Kisalu N, Kebela-Ilunga B, Mukaba T, Wright LL, Formenty P, Wolfe ND, Shongo RL, Tshioko F, Okitolonda E, et al. (2007). Endemic human monkeypox, Democratic Republic of Congo, 2001-2004. Emerg Infect Dis 13, 934–937. 2007/06/08.

2. Rimoin AW, Mulembakani PM, Johnston SC, Lloyd Smith JO, Kisalu NK, Kinkela TL, Blumberg S, Thomassen HA, Pike BL, Fair JN, et al. (2010). Major increase in human monkeypox incidence 30 years after smallpox vaccination campaigns cease in the Democratic Republic of Congo. Proc Natl Acad Sci U S A 107, 16262–16267. 2010/09/02.

3. Huang Y, Mu L and Wang W. (2022). Monkeypox: epidemiology, pathogenesis, treatment and prevention. Signal Transduct Target Ther 7, 373. 2022/11/03.

4. Yinka-Ogunleye A, Aruna O, Dalhat M, Ogoina D, McCollum A, Disu Y, Mamadu I, Akinpelu A, Ahmad A, Burga J, et al. (2019). Outbreak of human monkeypox in Nigeria in 2017-18: a clinical and epidemiological report. Lancet Infect Dis 19, 872–879. 2019/07/10.

5. Gross E. (2003). Update on emerging infections: news from the Centers for Disease Control and prevention. Update: Multistate outbreak of monkeypox--Illinois, Indiana, Kansas, Missouri, Ohio, and Wisconsin, 2003. Ann Emerg Med 42, 660–662; discussion 662-664. 2003/10/29.

6. Hendrickson RC, Wang C, Hatcher EL and Lefkowitz EJ. (2010). Orthopoxvirus genome evolution: the role of gene loss. Viruses 2, 1933–1967. 2011/10/14.

7. Grabenstein JD and Winkenwerder W, Jr. (2003). US military smallpox vaccination program experience. JAMA 289, 3278–3282. 2003/06/26.

8. Henderson DA, Inglesby TV, Bartlett JG, Ascher MS, Eitzen E, Jahrling PB, Hauer J, Layton M, McDade J, Osterholm MT, et al. (1999). Smallpox as a biological weapon: medical and public health management. Working Group on Civilian Biodefense. JAMA 281, 2127–2137. 1999/06/15.

9. Payne AB, Ray LC, Cole MM, Canning M, Houck K, Shah HJ, Farrar JL, Lewis NM, Fothergill A, White EB, et al. (2022). Reduced Risk for Mpox After Receipt of 1 or 2 Doses of JYNNEOS Vaccine Compared with Risk Among Unvaccinated Persons - 43 U.S. Jurisdictions, July 31-October 1, 2022. MMWR Morb Mortal Wkly Rep 71, 1560–1564. 2022/12/09.

10. Karem KL, Reynolds M, Hughes C, Braden Z, Nigam P, Crotty S, Glidewell J, Ahmed R, Amara R and Damon IK. (2007). Monkeypox-induced immunity and failure of childhood smallpox vaccination to provide complete protection. Clin Vaccine Immunol 14, 1318–1327. 2007/08/24.

11. Priyamvada L, Carson WC, Ortega E, Navarra T, Tran S, Smith TG, Pukuta E, Muyamuna E, Kabamba J, Nguete BU, et al. (2022). Serological responses to the MVA-based JYNNEOS monkeypox vaccine in a cohort of participants from the Democratic Republic of Congo. Vaccine 40, 7321–7327. 2022/11/08.

12. Townsend MB, Keckler MS, Patel N, Davies DH, Felgner P, Damon IK and Karem KL. (2013). Humoral immunity to smallpox vaccines and monkeypox virus challenge: proteomic assessment and clinical correlations. J Virol 87, 900–911. 2012/11/09.

13. Alzhanova D, Hammarlund E, Reed J, Meermeier E, Rawlings S, Ray CA, Edwards DM, Bimber B, Legasse A, Planer S, et al. (2014). T cell inactivation by poxviral B22 family proteins increases viral virulence. PLoS Pathog 10, e1004123. 2014/05/17.

14. Hammarlund E, Lewis MW, Hansen SG, Strelow LI, Nelson JA, Sexton GJ, Hanifin JM and Slifka MK. (2003). Duration of antiviral immunity after smallpox vaccination. Nat Med 9, 1131–1137. 2003/08/20.

15. Dubois ME, Hammarlund E and Slifka MK. (2012). Optimization of peptide-based ELISA for serological diagnostics: a retrospective study of human monkeypox infection. Vector Borne Zoonotic Dis 12, 400–409. 2012/01/06.

16. Kava CM, Rohraff DM, Wallace B, Mendoza-Alonzo JL, Currie DW, Munsey AE, Roth NM, Bryant-Genevier J, Kennedy JL, Weller DL, et al. (2022). Epidemiologic Features of the Monkeypox Outbreak and the Public Health Response - United States, May 17-October 6, 2022. MMWR Morb Mortal Wkly Rep 71, 1449–1456. 2022/11/11.

17. Mandrekar JN. (2010). Receiver operating characteristic curve in diagnostic test assessment. J Thorac Oncol 5, 1315–1316. 2010/08/26.

18. Crotty S, Felgner P, Davies H, Glidewell J, Villarreal L and Ahmed R. (2003). Cutting edge:long-term B cell memory in humans after smallpox vaccination. J Immunol 171, 4969–4973. 2003/11/11.

19. Hanna W and Baxby D. (2002). Studies in smallpox and vaccination. 1913. Rev Med Virol 12, 201–209. 2002/07/19.

20. Gilchuk I, Gilchuk P, Sapparapu G, Lampley R, Singh V, Kose N, Blum DL, Hughes LJ, Satheshkumar PS, Townsend MB, et al. (2016). Cross-Neutralizing and Protective Human Antibody Specificities to Poxvirus Infections. Cell 167, 684–694 e689. 2016/10/22.

21. Hammarlund E, Lewis MW, Carter SV, Amanna I, Hansen SG, Strelow LI, Wong SW, Yoshihara P, Hanifin JM and Slifka MK. (2005). Multiple diagnostic techniques identify previously vaccinated individuals with protective immunity against monkeypox. Nat Med 11, 1005–1011. 2005/08/09.

22. Earl PL, Americo JL, Wyatt LS, Eller LA, Whitbeck JC, Cohen GH, Eisenberg RJ, Hartmann CJ, Jackson DL, Kulesh DA, et al. (2004). Immunogenicity of a highly attenuated MVA smallpox vaccine and protection against monkeypox. Nature 428, 182–185. 2004/03/12.

23. Lane HC, Masur H, Edgar LC, Whalen G, Rook AH and Fauci AS. (1983). Abnormalities of B-cell activation and immunoregulation in patients with the acquired immunodeficiency syndrome. N Engl J Med 309, 453–458. 1983/08/25.

24. Shirai A, Cosentino M, Leitman-Klinman SF and Klinman DM. (1992). Human immunodeficiency virus infection induces both polyclonal and virus-specific B cell activation. J Clin Invest 89, 561–566. 1992/02/01.

25. Prigent J, Jarossay A, Planchais C, Eden C, Dufloo J, Kok A, Lorin V, Vratskikh O, Couderc T, Bruel T, et al. (2018). Conformational Plasticity in Broadly Neutralizing HIV-1 Antibodies Triggers Polyreactivity. Cell Rep 23, 2568–2581. 2018/05/31.

26. Duke-Cohan JS, Wollenick K, Witten EA, Seaman MS, Baden LR, Dolin R and Reinherz EL. (2009). The heterogeneity of human antibody responses to vaccinia virus revealed through use of focused protein arrays. Vaccine 27, 1154–1165. 2009/01/17.

27. Lantto J, Haahr Hansen M, Rasmussen SK, Steinaa L, Poulsen TR, Duggan J, Dennis M, Naylor I, Easterbrook L, Bregenholt S, et al. (2011). Capturing the natural diversity of the human antibody response against vaccinia virus. J Virol 85, 1820–1833. 2010/12/15.

28. Yefet R, Friedel N, Tamir H, Polonsky K, Mor M, Cherry-Mimran L, Taleb E, Hagin D, Sprecher E, Israely T, et al. (2023). Monkeypox infection elicits strong antibody and B cell response against A35R and H3L antigens. iScience 26, 105957. 2023/01/24.

29. Styer LM, Hoen R, Rock J, Yauney E, Nemeth K, Bievenue R and Parker MM. (2021). High-Throughput Multiplex SARS-CoV-2 IgG Microsphere Immunoassay for Dried Blood Spots: A Public Health Strategy for Enhanced Serosurvey Capacity. Microbiol Spectr 9, e0013421. 2021/07/29.

30. Reynolds SE and Moss B. (2015). Characterization of a large, proteolytically processed cowpox virus membrane glycoprotein conserved in most chordopoxviruses. Virology 483, 209–217. 2015/05/20.

31. Sereewit J, Lieberman NAP, Xie H, Bakhash S, Nunley BE, Chung B, Mills MG, Roychoudhury P and Greninger AL. (2022). ORF-Interrupting Mutations in Monkeypox Virus Genomes from Washington and Ohio, 2022. Viruses 14. 2022/11/12.

